# Antibody Responses After a Single Dose of ChAdOx1 nCoV-19 Vaccine in Healthcare Workers Previously Infected with SARS-CoV-2

**DOI:** 10.1101/2021.05.08.21256866

**Authors:** Sebastian Havervall, Ulrika Marking, Nina Greilert-Norin, Henry Ng, Ann-Christin Salomonsson, Cecilia Hellström, Elisa Pin, Kim Blom, Sara Mangsbo, Mia Phillipson, Jonas Klingström, Mikael Åberg, Sophia Hober, Peter Nilsson, Charlotte Thålin

## Abstract

**Background:** Recent reports demonstrate robust serological responses to a single dose of messenger RNA (mRNA) vaccines in individuals previously infected with SARS-CoV-2. Data on immune responses following a single-dose adenovirus-vectored vaccine expressing the SARS-CoV-2 spike protein (ChAdOx1 nCoV-19) in individuals with previous SARS-CoV-2 infection are however limited, and current guidelines recommend a two-dose regime regardless of preexisting immunity.

**Methods:** We compared spike-specific IgG and pseudo-neutralizing spike-ACE2 blocking antibodies against SARS-CoV-2 wild type and variants B.1.1.7, B.1.351, and P1 following two doses of the mRNA vaccine BNT162b2 and a single dose of the adenovector vaccine ChAdOx1 nCoV-19 in 232 healthcare workers with and without previous COVID-19.

**Findings:** The post-vaccine levels of spike-specific IgG and neutralizing antibodies against the SARS-CoV-2 wild type and all three variants of concern were similar or higher in participants receiving a single dose of ChAdOx1 nCoV-19 vaccine post SARS-CoV-2 infection (both < 11 months post infection (n=37) and ≥ 11 months infection (n=46)) compared to participants who received two doses of BNT162b2 vaccine (n=149).

**Interpretation:** Our data support that a single dose ChAdOx1 nCoV-19 vaccine serves as an effective immune booster after priming with natural SARS-CoV-2 infection up to at least 11 months post infection.

## Introduction

Recent reports demonstrate robust serological responses to a single dose of messenger RNA (mRNA) vaccines in individuals previously infected with SARS-CoV-2. These individuals develop spike-specific IgG and neutralizing antibody titers after one dose that are equivalent to or exceeding those observed after the second dose in vaccinees without preexisting immunity (1-7). In a study of 23 SARS-CoV-2 experienced individuals, a substantial enhancement of neutralizing antibody response against SARS-CoV-2 variants B.1.1.7 and B.1.351 by a single dose mRNA vaccine was noted (6). A second dose mRNA vaccine to individuals with prior SARS-CoV-2 infection add little value in terms of serological titers (1, 4, 7), and may be associated with increased risk of non-severe adverse events (8). Data on immune responses following a single-dose adenovirus-vectored vaccine expressing the SARS-CoV-2 spike protein such as ChAdOx1 nCoV-19 in individuals with previous SARS-CoV-2 infection are however limited. Current guidelines recommend a two-dose regime regardless of preexisting immunity (9).

We compared spike-specific IgG and their neutralizing capacity towards SARS-CoV-2 wild type and three variants of concern following two doses of the mRNA vaccine BNT162b2 and a single dose of the adenovector vaccine ChAdOx1 nCoV-19 in healthcare workers with and without previous COVID-19.

## Methods

The COMMUNITY (COVID-19 Biomarker and Immunity) study (10, 11) investigates long-term immunity after COVID-19 in 2149 healthcare workers at Danderyd Hospital, Stockholm, Sweden, included between April 15th and May 8th 2020. Blood samples are obtained every four months and SARS-CoV-2 spike-specific IgG are analyzed by multiplex antigen bead array (FlexMap3D, Luminex Corp) as previously described (10).

Between March 23rd and 31st, 2021, this sub study investigated serological responses following either two doses of BNT162b2 or one dose of ChAdOx1 nCoV-19 vaccination in 232 study participants with or without previous SARS-CoV-2 infection confirmed by seroconversion. Of these, 149 participants received two doses of BNT162b2 vaccine (36 previously infected participants with median age 52 (IQR 41-62), 86% female, and 113 SARS-CoV-2-naïve participants with median age 53 (IQR 42-58), 86 % women), and 83 participants received a single-dose ChAdOx1 nCoV-19 vaccine (46 participants ≥ 11 months post SARS-CoV-2 infection with median age 49 (IQR 41-60), 83% female, and 37 participants < 11 months post SARS-CoV-2 infection with median age 48 (IQR 40-54), 95 % women). 162 unvaccinated previously SARS-CoV-2 infected study participants with median age 49 (IQR 40-76), 93% female, and 80 unvaccinated seronegative study participants with median age 51 (IQR 41-58), 91% female, were enrolled as control groups.

Blood samples were collected at least two weeks post second dose BNT162b2 or at least one week post single dose ChAdOx1 nCoV-19 vaccination. Spike-specific IgG and their neutralizing capacity (analyzed by a pseudo-neutralizing spike-ACE2 assay) against SARS-CoV-2 wild type and variants B.1.1.7, B.1.351, and P1 were measured using the V-PLEX SARS-CoV-2 Panel 7 (IgG and ACE2, Meso Scale Diagnostics, USA) and expressed as arbitrary units (AU)/ml. For validation, spike-specific IgG antibodies against SARS-CoV-2 wild type were measured in the same samples using the multiplex antigen bead array (FlexMap3D, Luminex Corp) (10). Group comparisons were performed using Dunn’s Kruskal-Wallis multiple comparison test. The statistical significance threshold was set at 5%. Analyses were performed using GraphPad Prism 9.1.0 (GraphPad Software, Inc, USA). The study was approved by the Swedish Ethical Review Authority, and informed written consent was obtained from all participants.

## Results

The median levels of spike-specific IgG antibodies against the SARS-CoV-2 wild type and variants of concern B.1.1.7, B.1.351 and P1 were similar or higher in participants post SARS-CoV-2 infection (both < 11 months post infection and ≥ 11 months post infection) receiving a single dose ChAdOx1 nCoV-19 vaccine compared to participants who received two doses BNT162b2 vaccine (Figure 1 A-D, Figure S1). Neutralizing antibodies towards the SARS-CoV-2 wild type and all three variants reached higher levels in previously infected vaccinees receiving a single dose ChAdOx1 nCoV-19 vaccine than in participants fully vaccinated with BNT162b2 (Figure 2B-D).

**Figure 1.**
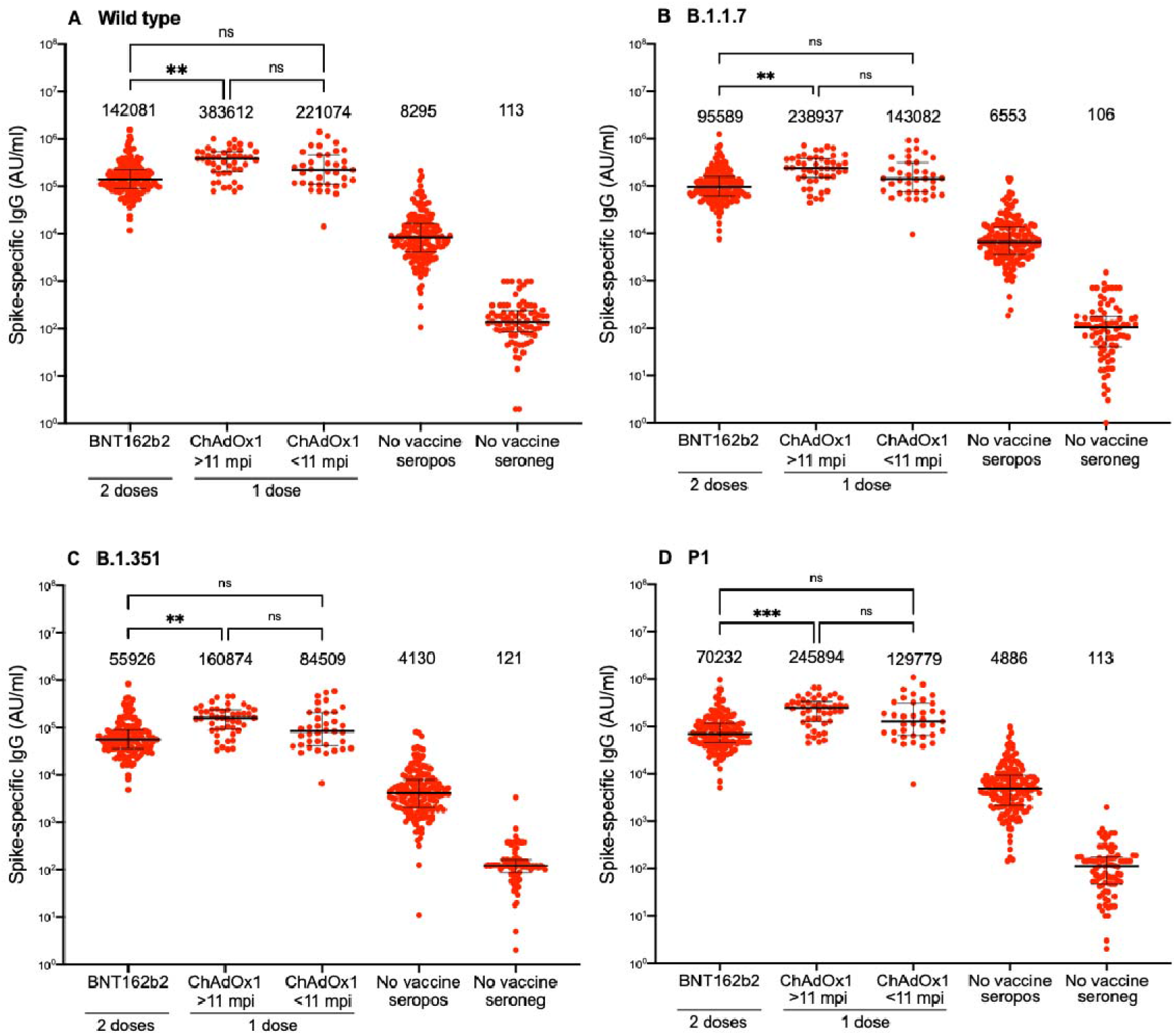
Spike-specific IgG responses against SARS-CoV-2 wild type and variants following two doses BNT162b2 in individuals with or without previous SARS-CoV-2 infection or one dose ChAdOx1 nCoV-19 vaccine in individuals with previous SARS-CoV-2 infection. A) Spike-specific IgG responses against wild type (A), B.1.1.7 (B), B.1.351 (C) and P1 (D). Numbers above each cluster indicate median AU/ml. Lines indicate medians with interquartile ranges. AU; arbitrary units, mpi; months post infection, ns; non-significant, *; p<0.05, **; p<0,01 ***; p<0.001, ****; p<0.0001.

**Figure 2.**
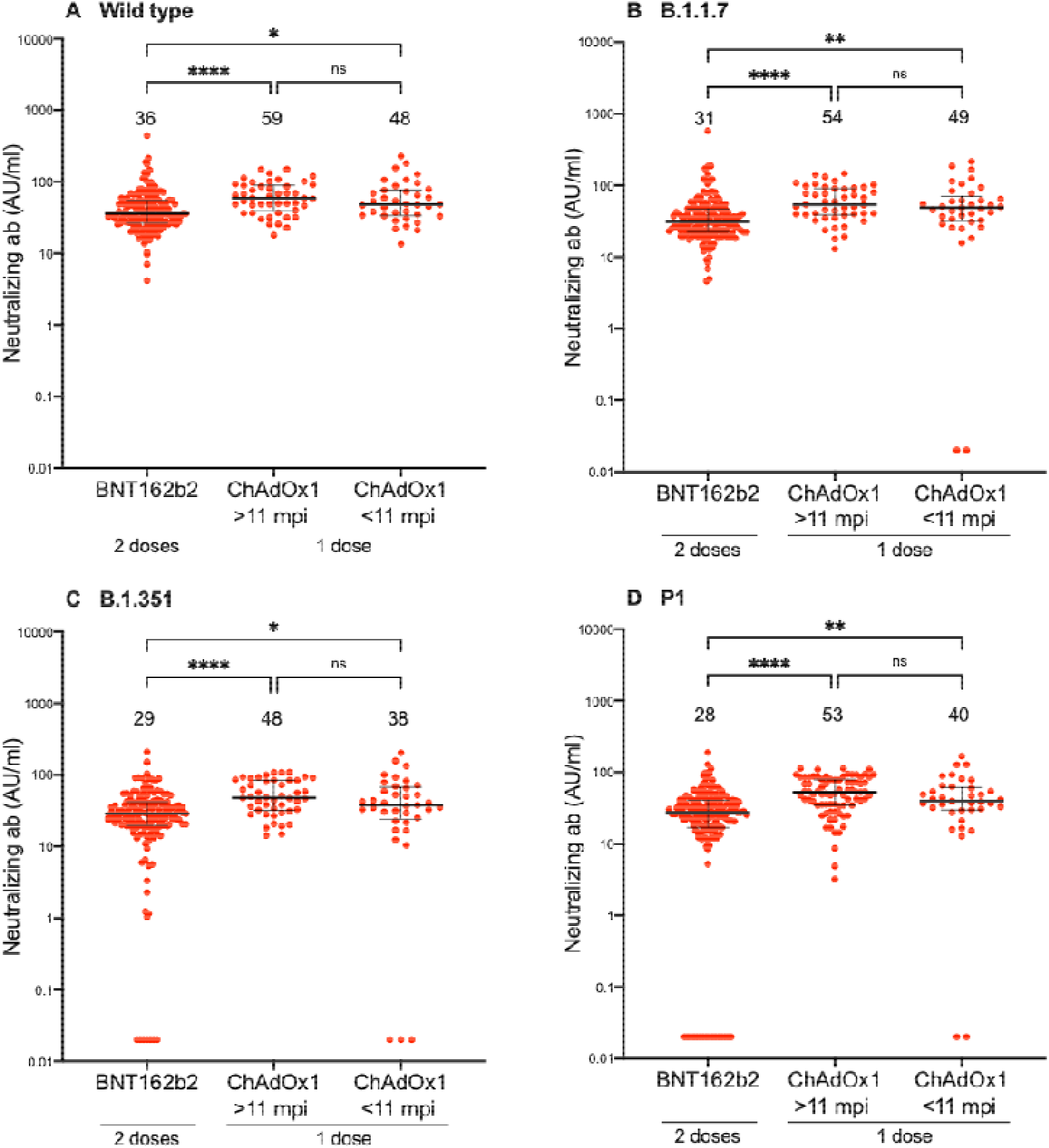
Neutralizing antibody responses against SARS-CoV-2 wild type and variants following two doses BNT162b2 in individuals with or without previous SARS-CoV-2 infection or one dose ChAdOx1 nCoV-19 vaccine in individuals with previous SARS-CoV-2 infection. Neutralizing antibody responses against wild type (A), B.1.1.7 (B), B.1.351 (C) and P1 (D). Numbers above each cluster indicate median AU/ml. Lines indicate medians with interquartile ranges. AU; arbitrary units, mpi; months post infection, ns; non-significant, *; p<0.05, **; p<0,01 ***; p<0.001, ****; p<0.0001.

## Discussion

A single dose of the adenovector vaccine ChAdOx1 nCoV-19 following natural infection elicited a robust serological response with broad neutralizing capacity against SARS-CoV-2 wild type and variants of concern. Neutralizing antibody levels exceeded those after two doses of the mRNA vaccine BNT162b2. Our data support that a single dose of adenovector vaccine serves as an efficacious immune booster after priming with natural SARS-CoV-2 infection up to at least 11 months post infection.

## Data Availability

Data is available upon reasonable request

## Acknowledgements

This work was funded by Jonas & Christina af Jochnick foundation; Lundblad family foundation; Region Stockholm; Knut and Alice Wallenberg foundation; Science for Life Laboratory (SciLifeLab); Erling-Persson family foundation.

## Author contribution

SeH, UM, PN and CT had full access to all of the data in the study and takes responsibility for the integrity of the data and the accuracy of the data analysis. SeH, UM, and CT designed the study, made statistical analysis, and drafted the manuscript. NG, HN, ACS, CH, EP, MÅ conducted the analysis and the administrative and material support. SeH, UM, KB, SM, JK, MP, MÅ, SoH, PN, CT interpreted the data, and all authors critically revised the manuscript. SoH, PN, CT obtained funding. MP, SoH, PN, CT supervised. All authors had final responsibility for the decision to submit for publication.

