## Supplemental Figure 1 for "Antibody Responses After a Single Dose of ChAdOx1 nCoV-19 Vaccine in Healthcare Workers Previously Infected with SARS-CoV-2"

**Supplementary**

**Figure 1S**. Spike IgG responses against SARS-CoV-2 wild type following two doses BNT162b2 in individuals with or without previous SARS-CoV-2 infection or one dose ChAdOx1 nCoV-19 vaccine in individuals with previous SARS-CoV-2 infection measured using the multiplex antigen bead array for validation. Numbers above each cluster indicate median AU. Lines indicate medians with interquartile ranges. AU; arbitrary units, mpi; months post infection, ns; non-significant.

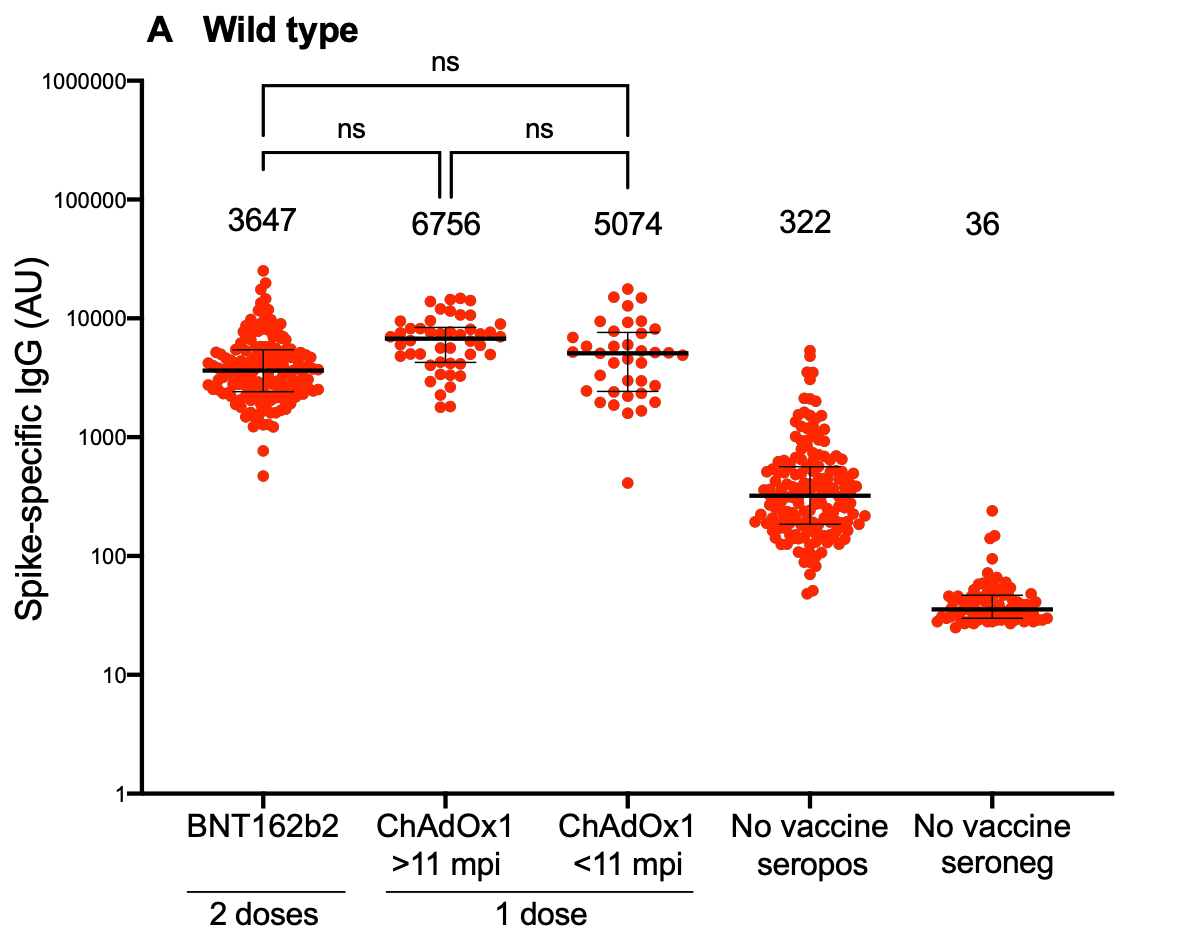
